# INCREASED REMOVAL SIGNALS ON ERYTHROCYTES OF ANEMIC CANCER PATIENTS

**DOI:** 10.64898/2026.06.23.26356301

**Authors:** Dimitris Matthaios, George Karatidis, Ioanna Balgkouranidou, Zoe Kyriakou, Konstantinos Anagnostopoulos, Charalampos Papadopoulos

**Affiliations:** Oncology Clinic, University Hospital of Alexandroupolis, Department of Medicine, Democritus University of Thrace, Alexandroupolis, Greece; Laboratory of Biochemistry, Department of Medicine, Democritus University of Thrace, Alexandroupolis, Greece

**Keywords:** cancer, anemia, Erythrocytes, CD47, MCP1, lactadherin, iron, Immunometabolism

## Abstract

**BACKGROUND:** Anemia is a negative factor in cancer, influencing the prognosis, quality of life and financial situation of cancer patients. Recent studies have shown that anemia in cancer is provoked by augmented erythrocyte removal.

**OBJECTIVE:** In this study we sought to investigate the molecular bases for erythrocyte removal in cancer patients with anemia. In particular, we explored the levels of erythrocyte CD47, lactadherin, calreticulin and MCP1.

**METHODS:** Thirty five anemic cancer patients (25 women, aged 66.4 ±11.35 years old) and twelve healthy non-anemic controls (8 men, aged 61.1±9.98 years old) participated in our study. Red blood cells were isolated throug multiple centrifugations, and were lysed with the use of Triton-X 100. The levels of CD47, lactadherin, calreticulin and monocyte chemoattrractant protein 1 were determined by ELISA.

**RESULTS:** Erythrocytes of anemic cancer patients display reduced CD47 (p<0.001), MCP1 (p=0.05), CD47 to lactadherin ratio (p<0.05), and increased lactadherin levels (p<0.01) in comparison to the healthy controls.

**DISCUSSION:** Reduced CD47 along with increased lactadherin possibly drive erythrocyte removal in anemic cancer patients.

**Conclusions:** The role of CD47 and increased lactadherin should be examined in the future as potential therapeutic targets and biomarkers for anemia diagnosis and iron dymsetabolism.

## INTRODUCTION

Cancer is the second leading cause of death worlwide[1]. A high percentage of cancer patients suffer from anemia, with the prevalence of anemia varying according to the type of cancer[2]. Anemia has been shown to reduce the life expectancy of cancer patients[3]. It also worsens the quality of life of cancer patients which is already reduced due to cancer[4]. Anemia also augments the financial cost of the cancer patients, with a large percentage of these patients being unable to cover the costs[5]. Anemia can also directly affect cancer progression and metastasis through the induction of immunosuppressive cells[6]. The current therapeutic protocol for cancer anemia involves the use of erythropoietin stimulating agents[7]. However, recent studies reveal adverse effect of erythropoietin on cancer cells[8,9].

The use of erythropoietin stimulating agents is based on the notion that cancer anemia is triggered by reduced erythrocyte production. However, a recent study in animal models showed that anemia is triggered by increased erythrocyte removal[10]. This event is provoked by the sytstemic inflammatory and metabolic environment caused by cancer[10]. In addition, a previous study had already shown that erythrocytes of anemic lung cancer patients exhibit increased phosphatidylserine exposure[11]. These studies prove that increased erythrocyte removal could serve as a therapeutic target in cancer anemia.

Erythrocyte removal is provoked by various mechanisms. Reduction of CD47, increase opsonization of exposed phosphatidylserine by lactadherin and calreticulin, increased chemokine release, reduced membrane fluidity and increased antibody and complement opsonization are the main mechanisms driving erythrocyte removal[12].

In this study we sought to investigate the molecular bases for erythrocyte removal in cancer patients with anemia. In particular, we explored the levels of CD47, lactadherin, calreticulin and MCP1 (monocyte chemoattrractant protein 1).

## METHODS

### PATIENTS

Thirty five anemic cancer patients (25 five women, aged 66.4 ±11.35 years old) and twelve healthy non-anemic controls (8 men, aged 61.1±9.98 years old) were recruited by the Oncology Clinic of University Hospital of Alexandroupolis.

### EXPERIMENTAL

#### Erythrocyte isolation

Blood samples (3 mL) were obtained from each individual, either patients or healthy controls, into tubes with 5.4 mg of EDTA. To isolate red blood cells, the blood was centrifuged at 200×g for 10 minutes at 4 °C, which separated the plasma and buffy coat. Next, 1 mL of the erythrocyte pellet was washed with cold saline and centrifuged again under identical conditions. The washing cycle was carried out four times, until a purified erythrocyte population was reached (13).

#### Lysis of erythrocytes

A volume of 1 mL of packed erythrocytes was lysed using Triton X-100 to achieve a final concentration of 0.1% (v/v) [14)].

#### Determination of the levels of CD47, MCP1, lactadherin and calreticulin

The levels of CD47, MCP1, lactadherin and calreticulin were determined by ELISA assay kits according to the manufacturer’s instructions (FineTest, China).

#### Statistical Analysis

Results are presented as mean ± standard deviation and median, unless otherwise indicated. Statistical analyses were conducted using R software version 4.0.29 [15]. Comparisons of group means employed Welch’s t-test [16], as it corrects for unequal variances and sample sizes, improving reliability, making it superior to Student’s t-test in scenarios like this research. The magnitude of differences was quantified using Glass’s Δ [17], appropriate when group standard deviations vary markedly. Effect size thresholds were defined as Δ < 0.2 (very small), 0.2–0.41 (small), 0.41–0.63 (moderate), and ≥0.63 (large). Results are presented as t (degrees of freedom) = test statistic, p-value, Glass’s Δ with its 95% confidence interval, statistical power (1−β) at a significance level of α = 0.05, and the 95% confidence interval for the difference in means between healthy and ancer anemia groups, as summarized in Table 1.

**Table 1:**
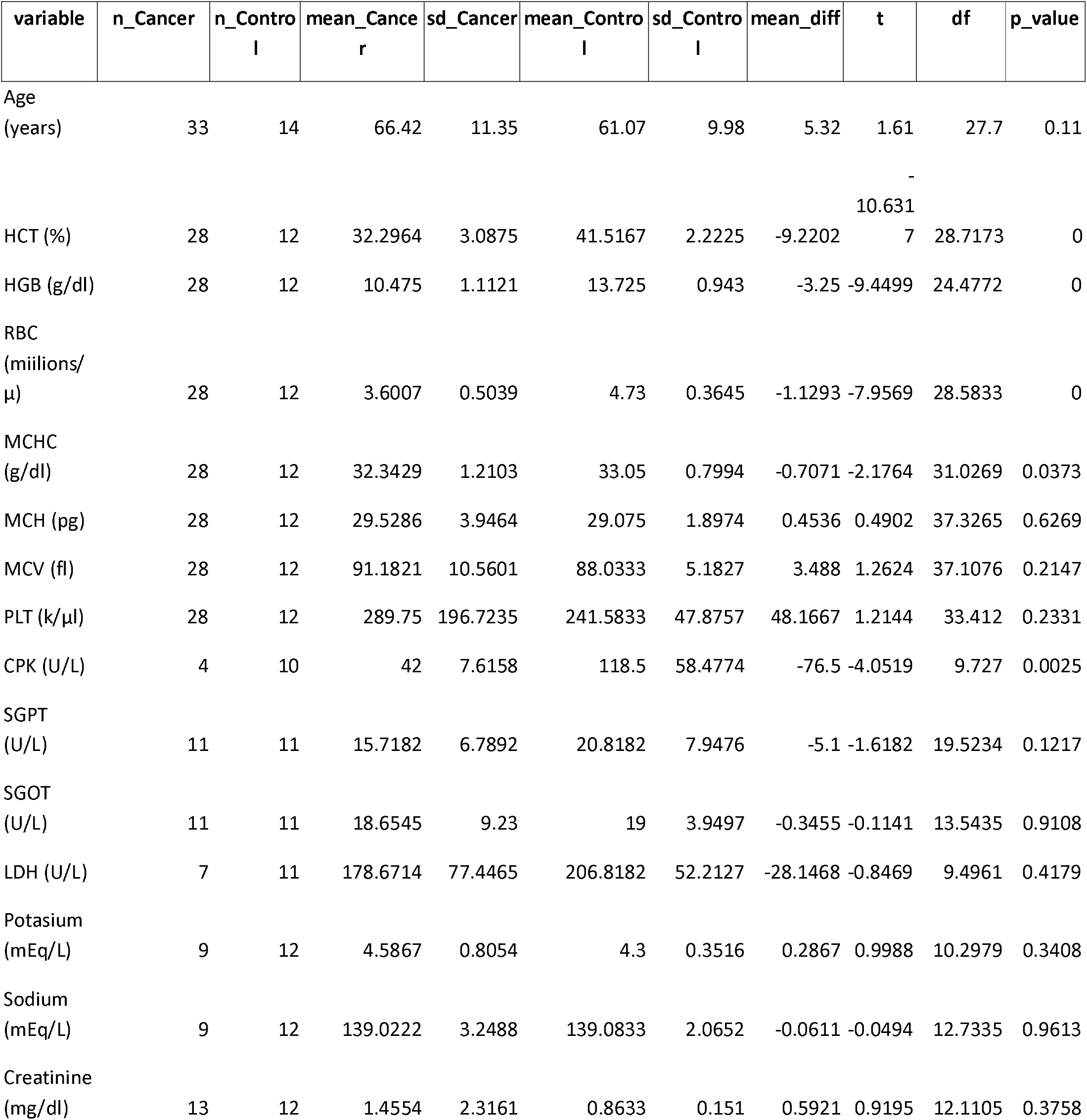
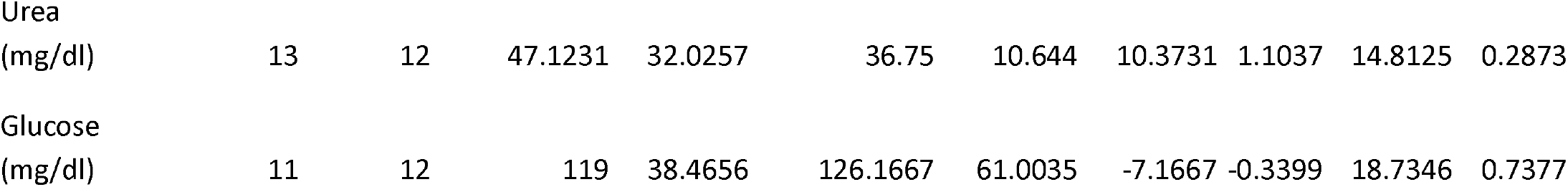
Anthropometric characteristics of anemic cancer patients and healthy controls. CPK: creatine kinase; HCT: hematocrite; HGB: hemoglobin mass; LDH: lactate dehydrogenase; MCH: mean corpuscular hemoglobin; MCHC: Mean corpuscular hemoglobin concentration; MCV: mean corpuscular volume; PLT: platelets; RBC: red blood cell count; SGPT: alanine transaminase; SGOT: aspartate transaminase

To explore potential biological links, we performed correlation analyses between the blood markers of interest (HCT, HGB, RBC, MCHC) and molecular features (CD47 to Lactadherin Ratio,, CD47, MCP1, Calreticulin, Lactadherin). We estimated Pearson’s correlation (r) for linear associations (with 95% CI via Fisher’s z) and Spearman’s rank correlation (ρ) for monotonic, outlier robust associations. Because group membership can induce spurious associations, we additionally computed partial Pearson correlations adjusted for Group (Cancer vs Control) by residualizing each variable on Group and correlating the residuals; corresponding test statistics, degrees of freedom, and p-values were reported. FDR control using BH was applied separately to each correlation family (Pearson, Spearman, partial), given 28 planned pairs per method.

## RESULTS

In the group comparison (Cancer vs. Control), statistically significantly lower HCT, HGB, RBC and CPK values were observed in the Cancer group, with very large effect sizes for the complete blood count indices (Hedges’ g≈−3 to −2.4).

Erythrocytes of anemic cancer patiens exhibit reduced CD47 content in comparison to the healthy controls (p<0.001). Lactadherin levels were also reduced in the erythrocytes of anemic cancer patients compared to the healthy controls(p<0.01). Interestingly, the ratio of CD47 to lactadherin ratio, a marker of the balance between the anti phagocytosis and prophagocytosis signals, is reduced in patient erythrocytes (p<0.05).

The levels of the chemokine MCP1 were found reduced in the erythrocytes of anemic cancer patients in comparison to the healthy controls (p=0.05). This likely signifies increased MCP1 release from the erythrocytes of anemic cancer patients.

The levels of calreticulin were not different between patients and controls.

All results can be found in figure 1.

**Figure 1:**
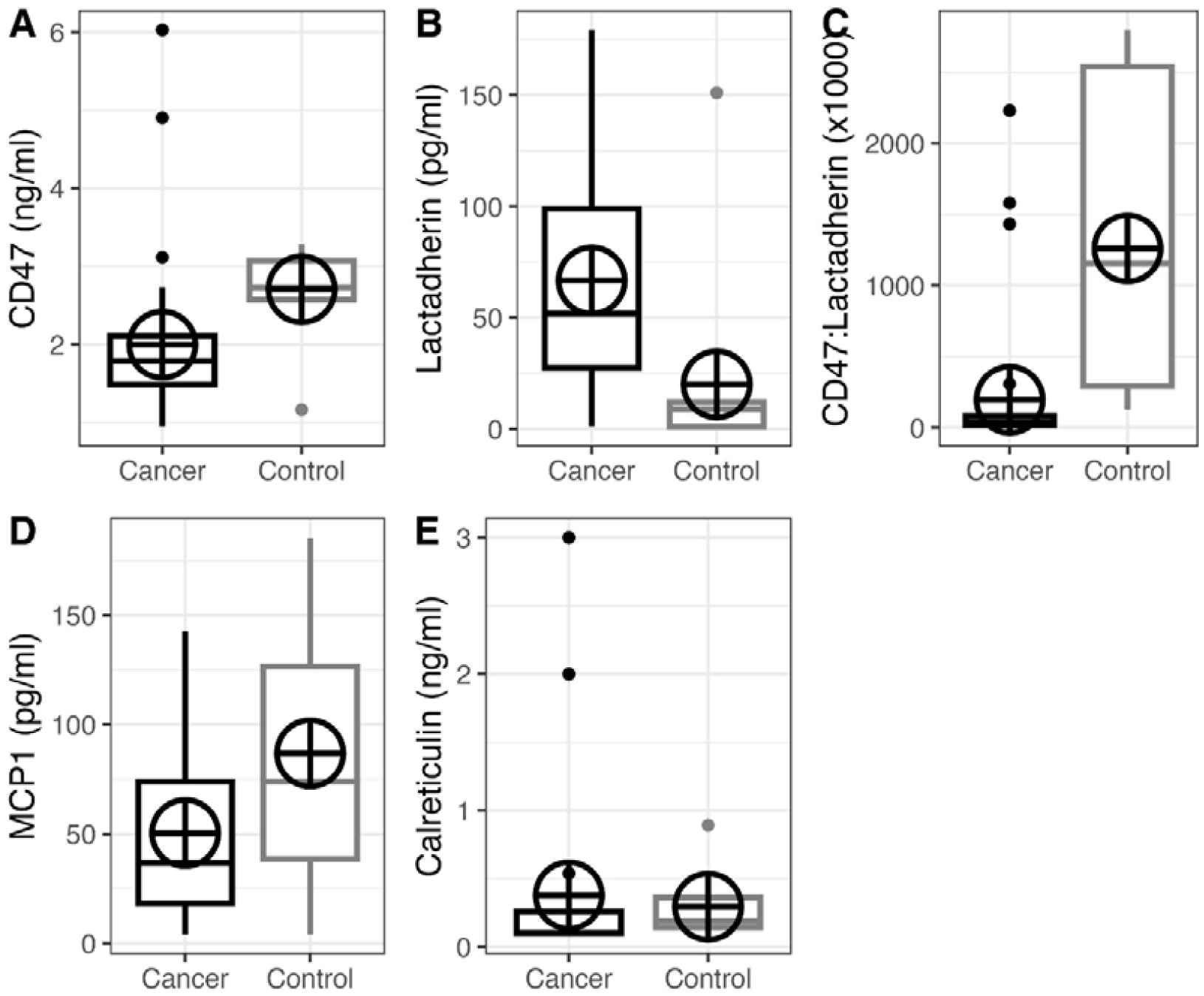
A. CD47 concentration on erythrocytes of anemic cancer patients (n=35) and healthy controls (n=12) , p<0.001. B. Lactadherin concentration on erythrocytes of cancer patients (35) and healthy controls (n=11), p<0.01. C. CD47 to lactadherin ratio on erythrocytes of anemic cancer patients (n=35) and healthy controls (n=9), p<0.05. D. MCP1 concentration on erythrocytes of anemic cancer patients (n=35) and healthy controls (n=14), p=0.05. E. Calreticulin concentration on erythrocytes of anemic cancer patients (n=35) and healthy controls (n=12), p=0.56. CD47: cluster differentiation 47; MCP1: monocyte chemoattrractant protein 1.

In the correlation analyses, hematocrite and hemoglobin mass showed a consistent positive correlation with CD47 and the indices Cd47 to Lactadherin ratio (CD47/Lactadherin), while hematocrite was positively correlated with MCP1. Conversely, Lactadherin was negatively correlated with red blood cell count, hemoglobin mass and hematocrite (supplementary material).

However, after adjustment for Group (partial Pearson), no correlations retained statistical significance against the FDR, suggesting that the differences between Cancer/Control explain a significant part of the unadjusted findings. Multiple tests were controlled with FDR (Benjamini– Hochberg) and pairwise deletion was applied for missing values.

## DISCUSSION

Anemia is a negative factor in cancer, influencing the prognosis, quality of life and financial situation of cancer patients[1-5]. Recent studies have shown that anemia in cancer is provoked by augmented erythrocyte removal [10,11]. Unravelling the molecular signals that drive erythrocyte removal could lead to the discovery of novel therapeutic targets and biomarkers.

We report reduced CD47 content in the erythrocytes of anemic cancer patients. This reduction can lead to the reduction of the main anti-phagocytosis signal, facilitating erythrophagocytosis. We also reported, augmented lactadherin levels on erythrocytes of patients. This result possibly signifies the recognition of exposed phosphatidylserine, creating a pro-phagocytosis sighal.

Importantly, the ratio of CD47 to lactadherin was decreased. This is of major importance because erythrophagocytosis is determined by the balance between the prophagocytosis and the antiphagocytosis signals. Our results indicate that the CD47 to lactadherin ratio could serve as an early anemia biomarker in cancer patients. In addition, this ratio could serve as a therapeutic target to reduce erythrocyte removal and prevent anemia.

Regarding the molecular determinants for the reduction of the CD47 to lactadherin ratio, although not examined in our study, a previous report indicated that cancer-induced systemic metabolic changes drive erythrocyte removal[10]. Hence, the systemic and local immunometabolic landscape during cancer could drive erythrocyte removal (10,18)

The effect of erythrophagocytosis on iron metabolism is also of major importance. Cancer cells acquire higher levels of iron, which is mainly derived by tumor macrophage – mediated erythrophagocytosis and subsequent iron release to the tumor microenvironment [19]. Hence, we suggest that erythrophagocytosis could serve as a pathogenic mechanism in cancer growth through iron Immunometabolism.

We also report reduced MCP1 content in the erythrocytes of the patients, albeit not statistically significant. Accordingly, it is of major importance that reduced chemokine scavenging ability of erythrocytes has been found to drive cancer progression in prostate cancer patients[20]. Although we do not provide mechanistic proof, we presume this could be a possible mechanism in the cancer environment.

## Conclusions

Overall, our study indicates decreased erythrocyte CD47 to lactadherin ratio in anemic cancer patients, implying that at least in part, anemia is caused by increased recognition of erythrocytes by the immune system. The impact on anemia and iron Immunometabolism should be further investigated in the future.

## Data Availability

All data produced in the present study are available upon reasonable request to the authors

## ABBREVIATION

CD47: cluster differentiation 47
CPK: creatine kinase
HCT: hematocrite
HGB: hemoglobin mass
LDH: lactate dehydrogenase
MCH: mean corpuscular hemoglobin
MCHC: Mean corpuscular hemoglobin concentration
MCP1: monocyte chemoattrractant protein 1
MCV: mean corpuscular volume
PLT: platelets
RBC: red blood cell count
SGPT: alanine transaminase
SGOT: aspartate transaminase

## AUTHORS’ CONTRIBUTIONS

The authors confirm their contribution to the paper as follows: study conception and design: CP; data collection: GK, ZK, IB; Data Analysis or Interpretation: KA; methodology: CP, DM, KA; investigation: CP, DM. All authors reviewed the results and approved the final version of the manuscript.

## ETHICAL APPROVAL AND CONSENT TO PARTICIPATE

The Scientific Council of the University Hospital of Alexandroupolis and the Ethics Committee approved the study (protocol code EΣ12/Θ11/08-05-2025: and approval date 08/05/2025). Every participant provided a written informed consent.

## HUMANS AND ANIMALS RIGHT

No animals were used in this research. All procedures performed in studies involving human participants were in accordance with the ethical standards of institutional and/or research committees and with the 1975 Declaration of Helsinki, as revised in 2013.

## CONSENT FOR PUBLICATION

Informed consent was obtained from the participants.

## STANDARDS OF REPORTING

Strobe guidelines were followed.

## AVAILABILITY OF DATA AND MATERIALS

The data supporting the findings of the article is available within the article.

## ACKNOWLEDGEMENTS

Declared none.

